# Evaluating the Potential of AI-Generated Synthetic Diaries in Parkinson’s Disease Research

**DOI:** 10.1101/2025.05.02.25326887

**Authors:** Farhan Raza, Shahryar Wasif, Dhruvil Patel, Taylor Chomiak, Bin Hu

**Affiliations:** Canadian Open Digital Health (OpenDH) program, University of Calgary, Calgary, Alberta, Canada T2N 4N1; Biomedical Sciences, Cumming School of Medicine, University of Calgary, Calgary, AB T2N 4N1, Canada; Department of Psychology, Faculty of Arts, University of Calgary, Calgary, AB T2N 4N1, Canada; Division of Translational Neuroscience, Department of Clinical Neurosciences, Hotchkiss Brain Institute, University of Calgary, Calgary, AB T2N 4N1, Canada

**Keywords:** Artificial Intelligence, Large language models, GPT-4o, Parkinson’s disease, Synthetic data, Patient diaries, Data privacy

## Abstract

The integration of Artificial Intelligence (AI), particularly large language models like GPT-4o, into Parkinson’s Disease (PD) research presents a novel approach for generating synthetic patient diaries. These technologies offer potential benefits, including addressing data privacy concerns, overcoming limited sample sizes, and accelerating research timelines by providing alternative data sources. By leveraging its internal knowledge, GPT-4o demonstrated the capability to replicate overall symptom prevalence distributions observed in a real PD patient dataset without significant statistical deviation.

Despite these advantages, the widespread utility of AI-generated diaries based solely on internal knowledge is hindered by significant limitations identified in this case study. Key challenges include the failure to capture complex inter-variable correlations essential for understanding symptom co-occurrence, and a lack of the narrative richness, contextual depth, and linguistic nuance found in authentic patient reports. These findings underscore the constraints of current models in replicating real-world patient experiences without specific domain grounding. Addressing these challenges requires a multifaceted approach, including domain-specific fine-tuning, enhanced prompt engineering, and potentially hybrid data strategies to improve fidelity for high-stakes research applications. This case study explored the baseline capabilities and limitations of using GPT-4o’s internal knowledge for synthetic PD diary generation. It emphasizes the need for a balanced approach, acknowledging the potential for exploratory uses while highlighting the necessity for rigorous validation and further development before deployment in contexts requiring high fidelity. By fostering continued research and methodological refinement, AI-driven synthetic data generation can be better harnessed to support PD research and ultimately improve patient understanding.

## Introduction

Parkinson’s disease (PD) stands as the most prevalent neurodegenerative movement disorder, impacting millions globally [3]. Primarily characterized by motor symptoms like tremor, rigidity, and bradykinesia, PD also encompasses a wide array of non-motor symptoms that significantly diminish quality of life [3]. The underlying pathology involves the progressive degeneration of dopaminergic neurons in the substantia nigra and the aggregation of misfolded α-synuclein in Lewy bodies [3]. Despite advances in symptomatic treatments, such as dopamine replacement therapy, current interventions do not arrest disease progression [3].

The global prevalence of PD is escalating, driven by aging populations, with estimates suggesting up to 1% prevalence in individuals over 60 [3]. The disease levies a profound emotional, social, and economic toll on patients, caregivers, and healthcare systems. This escalating impact underscores the critical need for innovative methodologies to propel PD research and enhance patient outcomes.

### Parkinson’s Disease Diaries

Patient diaries are a long-established tool in PD research and clinical management, offering vital insights into symptom fluctuations, treatment responses, and overall quality of life. These self-reported instruments enable patients to chronicle changes in both motor and non-motor symptoms, aiding clinicians and researchers in comprehending the temporal dynamics of disease progression and treatment effectiveness [19].

Diaries prove particularly valuable for capturing complex phenomena like “on” and “off” states or dyskinesias [15]. An “on” state signifies periods of good symptom control, typically medication-induced, facilitating smoother movement. Conversely, an “off” state emerges as medication effects wane, leading to symptom recurrence or worsening [24]. The transient and variable nature of these states makes them challenging to assess accurately during brief clinical encounters. Real-time diary recording provides a more comprehensive representation of daily experiences, deepening understanding of treatment efficacy and motor fluctuation challenges [24]. For instance, the PD home diary, pioneered by Hauser et al. [15], is widely used in clinical trials to quantify motor fluctuations by categorizing patient states (“on without dyskinesia,” “on with dyskinesia,” “off”), providing objective therapeutic assessment endpoints [24]. Furthermore, diaries offer qualitative insights into the daily impact of PD, enriching the understanding of disease burden [19].

### Limitations of Diaries in Research

Despite their value, patient diaries pose significant research challenges, primarily concerning privacy and data management. Diaries often contain sensitive personal health information requiring stringent protection under privacy regulations. While research on diary privacy often focuses on contexts like intensive care units (ICUs), the principles are directly applicable to PD diaries [20, 28]. Anonymizing and securing such data is time-consuming and costly. Logistical hurdles, including patient burden and data management complexities, can impede implementation and delay research, particularly noted in fields like oncology and in resource-limited settings [10, 22]. The COVID-19 pandemic further highlighted how anonymity and confidentiality requirements can complicate data collection processes [5].

### Using AI Synthetic Diaries to Overcome Challenges

Generating synthetic patient diaries using Artificial Intelligence (AI) presents a promising avenue to circumvent these limitations. By creating realistic yet artificial datasets, AI can address privacy issues inherent in using real patient data. Advanced large language models (LLMs), such as GPT-4o, possess the potential to generate synthetic entries that closely mirror real-world data, capturing key aspects of symptoms, experiences, and treatment responses [9, 27]. This approach not only safeguards privacy but also enables researchers to work with larger, tailored datasets, addressing sample size constraints and facilitating more comprehensive studies within shorter timeframes [7].

### Large Language Models as a Potential Solution

Traditional synthetic data generation methods include rule-based systems and machine learning models like Generative Adversarial Networks (GANs). Rule-based systems, like Synthea [21], use predefined logic based on expert knowledge, proving effective for structured data but potentially struggling with the nuance of unstructured diary entries [8]. GANs learn underlying data distributions to generate realistic outputs [12] but face challenges with PD diaries, requiring large, high-quality, domain-specific datasets (often unavailable due to privacy concerns) and risking issues like mode collapse, limiting output diversity [6].

While GANs have seen some use in PD for specific data types (e.g., speech), their application to comprehensive diaries is limited due to complexity and subjectivity [18]. LLMs like GPT-4o offer a compelling alternative due to their advanced natural language processing capabilities trained on diverse text [2, 27]. Their proficiency in generating coherent, contextually relevant text suggests suitability for simulating the intricate narratives within patient diaries, potentially capturing subtleties of symptom progression, treatment effects, and daily life impacts [27].

Leveraging the internal knowledge encoded within LLMs allows synthetic diary generation without direct reliance on sensitive patient data, mitigating privacy risks [16]. Furthermore, LLM scalability enables the creation of large, diverse datasets, addressing limitations of sample size and data variability [7]. This approach promises to enhance the realism and utility of synthetic data, accelerating PD research by providing readily available, high-quality datasets.

### Objective of Study

This study aims to evaluate the feasibility, accuracy, and efficacy of using GPT-4o to generate synthetic patient diaries for PD research based *solely* on its internal knowledge, without fine-tuning on real PD diary data. Specifically, we sought to determine how well AI-generated diaries replicate real-world patient-reported symptom patterns, activities, and qualitative experiences. By comparing synthetic diaries to a real patient dataset using statistical and qualitative analyses, this study assesses whether AI can mitigate key challenges in diary-based research (privacy, sample size, data collection time). The findings aim to inform the potential utility and limitations of using AI-generated data as a reliable and ethical resource for advancing PD research.

## Methods

### Data Sources and Study Design

This study evaluated the capability of GPT-4o, OpenAI’s large language model, to generate synthetic diary data relevant to Parkinson’s disease research. The reference dataset consisted of records from 57 individuals diagnosed with Parkinson’s disease who were participants in registered clinical trials evaluating Ambulosono music based brisk walking exercise interventions (e.g., ISRCTN06023392) [111, 112]. Crucially, these records were fully anonymized prior to their inclusion in this evaluation study, ensuring individual patient identities were protected and adhering to strict ethical guidelines with Ethical approval obtained from the University of Calgary Conjoint Health Research Ethics Board. The reference “real” dataset comprised anonymized diary entries, including demographics (age, gender, education, disease duration), qualitative daily experience comments, physical activity logs, and responses to 12 predefined symptom questions (e.g., dyskinesia, freezing of gait, falls). The real dataset underwent preprocessing to standardize formats for analysis.

Management of patient data privacy was a central consideration throughout this research, as evidenced by the methodological design. The ‘real’ dataset utilized for comparative analysis consisted of records that were fully anonymized prior to their inclusion in this study, ensuring individual patient identities were protected. Critically, the generation of the synthetic dataset deliberately circumvented the use of extensive sensitive patient information; instead, it relied on GPT-4o’s internal knowledge, guided only by the aggregate demographic distribution and a single, non-identifiable structural example from the real data. Furthermore, the study utilized a GPT-4o enterprise account, under terms which explicitly forbid OpenAI from using the provided input data for their own model training, adding an additional layer of data security during the synthetic data generation process. This overall approach directly addresses the inherent privacy challenges associated with traditional patient diary research and underscores the study’s commitment to handling patient information responsibly and securely.

### Synthetic Data Generation

The synthetic dataset was generated by prompting GPT-4o with the demographic distribution of the real dataset and a single real diary entry as a structural template. This template included: (1) qualitative comments (e.g., “Today was a bit challenging; I felt tired but managed to take a short walk around the house”); (2) an activity log example (e.g., “15 minutes of stretching exercises”); and (3) the format for 12 symptom-related questions with example responses (e.g., “Do you experience freezing of gait? Sometimes”). Using this template and its internal knowledge of Parkinson’s disease, GPT-4o generated 500 synthetic diary entries for hypothetical patients matching the real dataset’s demographics. Each synthetic entry included qualitative comments, activity logs, and responses to the 12 symptom questions. Quality control checks verified adherence to demographic specifications and the sample format.

### Evaluation Framework

A multi-faceted framework involving thematic, narrative, semantic, and statistical analyses was employed to compare the synthetic and real datasets. Qualitative approaches examined thematic content and emotional tone in open-ended comments, while quantitative methods assessed linguistic features and statistical distributions.

### Thematic Analysis

Thematic analysis identified recurring patterns separately within each dataset. Open coding utilized Natural Language Processing (NLP), specifically Term Frequency-Inverse Document Frequency (TF-IDF) vectorization, to highlight key terms. KMeans clustering (k=5) grouped entries based on these vectorized patterns. Axial coding organized clusters into broader categories focusing on semantic relationships (e.g., emotional language, symptom tracking). Selective coding connected these categories to overarching themes, such as “Coping and Resilience” (synthetic) vs. “Structured Health Monitoring” (real).

### Narrative Analysis

Narrative analysis explored storytelling structure, coherence, detail, temporal markers, and subjective vs. objective observations. Representative entries were selected from both datasets to illustrate key differences in narrative complexity and emotional depth.

### Quantitative Content Analysis

OpenAI advanced data analytics comprising Python scripts and NLP tools which provided quantitative metrics for Word Diversity or Ratio of unique words to total sentences (via regex tokenization); Sentence Complexity or Average words per sentence; Emotional Prevalence or Frequency of emotional terms using an expanded NRC Emotion Lexicon (accounting for tense variations); Active/Passive Voice Ratio or Proportion of passive markers (“was,” “were,” “had been”) relative to total words. OpenAI’s GPT-4o assisted in identifying broader linguistic patterns and keywords.

Closed-ended symptom responses were analyzed for clinical relevance and statistical similarity. We used Chi-squared tests to compare frequency distributions of the 12 predefined symptoms between datasets. Spearman’s correlation coefficients were computed for symptom variables within each dataset (using pairwise deletion for missing real data). Correlation matrices were generated, and upper-triangular elements were flattened into vectors. Similarity was quantified using mean absolute difference, root mean squared difference, and correlation of correlations.

## Results

### Descriptive Statistics

Descriptive statistics for the real dataset (Table 1) and synthetic dataset (Table 2) reveal key differences. The real dataset had limited observations (n=1 to 16 per variable), whereas the synthetic dataset provided 500 observations per variable. Notably, Bed Mobility (BED) and Falls (Fal) responses were absent in the real data subset analyzed, though tracked, but present in the synthetic data, suggesting GPT-4o simulated a broader range of potential patient experiences.

**Table 1.** Descriptive statistics for the real dataset derived from symptom diaries, summarizing how often symptoms were reported and their mean values across variables: Wearing Off (WO), Dyskinesia (DY), Anxiety (AX), Freezing of Gait (FOG), Lack of Confidence (LOC), Imbalance (IB), Near Falls (NFal). Cognitive Memory (CM), and Sickness (SK). Measures include counts, means, standard deviations, and ranges (minimum, maximum, and quartiles). The data reflects small sample sizes (n = 1 to 16) and limited variability in some symptoms, such as FOG and LOC, which were consistently rated at their maximum values.

| *Statistic* | *WO* | *DY* | *AX* | *FOG* | *LOC* | *IB* | *NFal* | *CM* | *BED* | *Fal* | *SK* |
| --- | --- | --- | --- | --- | --- | --- | --- | --- | --- | --- | --- |
| count | 12 | 8 | 8 | 2 | 2 | 10 | 9 | 16 | 0 | 0 | 1 |
| mean | 1.67 | 1.50 | 1.50 | 3 | 3 | 1.40 | 1.44 | 2.81 | N/A | N/A | 1.00 |
| std | 0.78 | 0.93 | 0.93 | 0 | 0 | 0.84 | 0.88 | 0.40 | N/A | N/A | N/A |
| min | 1 | 1 | 1 | 3 | 3 | 1 | 1 | 2 | N/A | N/A | 1 |
| 25% | 1 | 1 | 1 | 3 | 3 | 1 | 1 | 3 | N/A | N/A | 1 |
| 50% | 1.5 | 1 | 1 | 3 | 3 | 1 | 1 | 3 | N/A | N/A | 1 |
| 75% | 2 | 1.5 | 1.5 | 3 | 3 | 1 | 1 | 3 | N/A | N/A | 1 |
| max | 3 | 3 | 3 | 3 | 3 | 3 | 3 | 3 | N/A | N/A | 1 |

\*Variables BED (Bed Mobility) and Fal (Falls) were not observed in the real dataset but were available for tracking

**Table 2.**
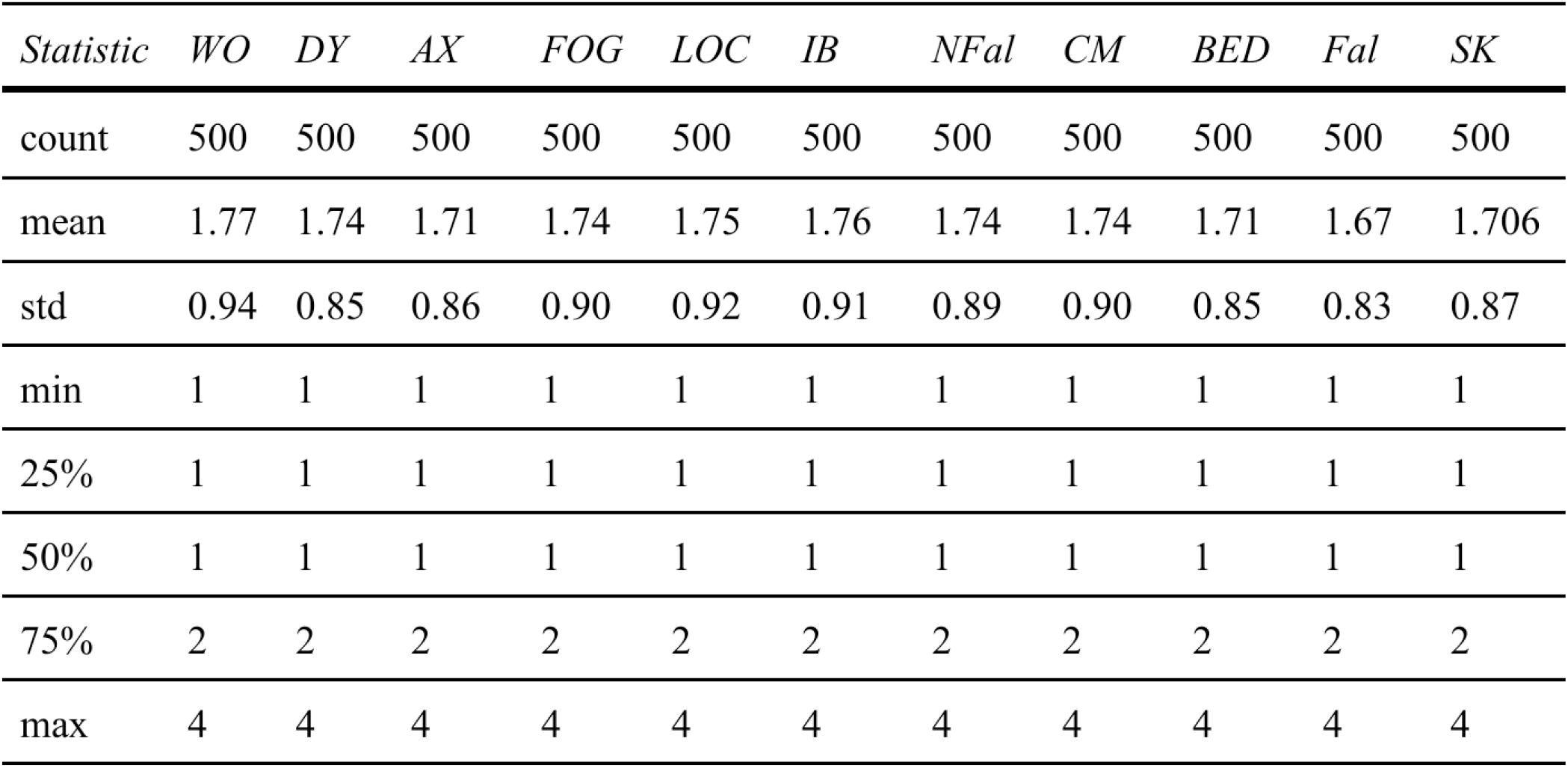
Descriptive statistics for the synthetic dataset generated to simulate symptom diary data. Variables include Wearing Off (WO), Dyskinesia (DY), Anxiety (AX), Freezing of Gait (FOG), Lack of Confidence (LOC), Imbalance (IB), Near Falls (NFal), Cognitive Memory (CM), Bed Mobility (BED), Falls (Fal), and Sickness (SK). The synthetic data provides measures of central tendency (mean, median), dispersion (standard deviation), and range (minimum, maximum, and quartiles) for 500 simulated entries per variable.

Mean symptom scores were generally higher in the synthetic data (e.g., WO mean: 1.77 synthetic vs. 1.67 real). Standard deviations were relatively consistent across synthetic variables (0.83-0.94) but more variable in the real dataset (0-0.93). Interquartile ranges (IQRs) were uniform in synthetic data (typically 1-2) but narrower for some real variables (e.g., FOG, LOC, SK showed no variability), reflecting the small sample size and limited observed range (max 3) in the real data compared to the broader range (max 4) generated synthetically. These differences highlight the synthetic dataset’s greater simulated variability versus the constraints of the small real sample.

*Code and prompts are available from corresponding authors upon reasonable request*.

### Symptom Prevalence

Chi-squared tests comparing the frequency distributions of the 12 predefined symptom responses revealed no statistically significant differences between the real and synthetic datasets (p > .05 for all symptoms) (Table 3). For instance, freezing of gait (χ^2^ = 0.01, p = 0.92) and dyskinesia (χ^2^ = 3.21, p = 0.78) showed comparable prevalence. This indicates GPT-4o, based on its internal knowledge, successfully replicated the overall distribution patterns of reported symptoms found in the real data sample without significant deviation.

Table 3. Chi-Square Test Results comparing the distribution of categorical variables between Synthetic Parkinson’s Data and Legacy Data… *(Content as in original)*

### Evaluation of Between-Symptom Relationships

Spearman correlation matrices were generated for both datasets to assess the replication of symptom co-occurrence patterns (Figures 1 & 2). The real dataset (Figure 1) showed several moderate-to-high correlations (e.g., WO and AX, r = 0.80), though many correlations were missing (NaN) due to low variance or non-overlapping observations for variables like FOG, LOC, CM, and SK.

**Figure 1.**
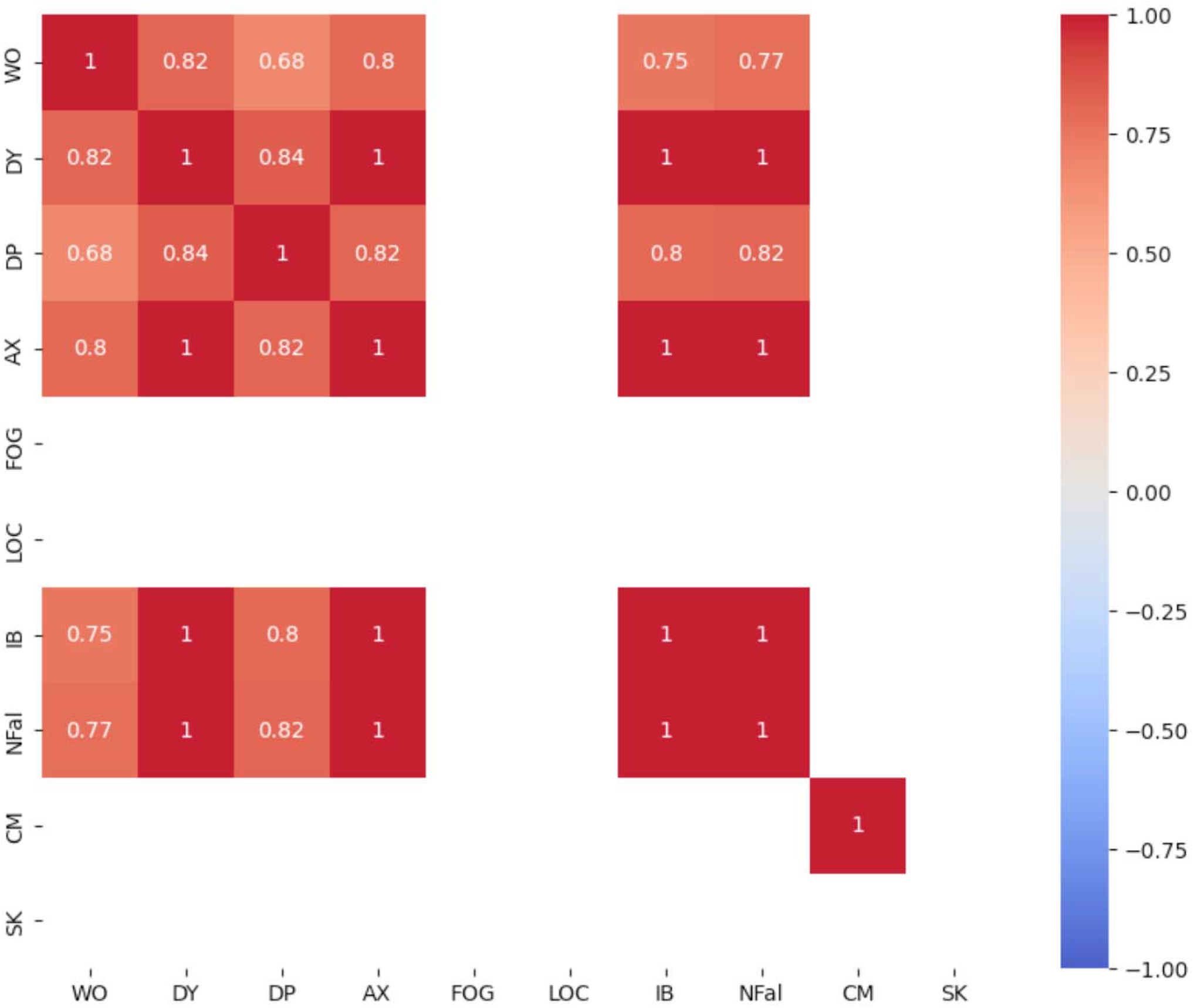
Spearman correlation matrix for symptom frequency variables in the real dataset. This heatmap depicts pairwise Spearman correlation coefficients (r) for ten symptom variables (WO, DY, DP, AX, FOG, LOC, IB, NFal, CM, SK), with values ranging from -1 (perfect negative correlation) to 1 (perfect positive correlation). Strong positive relationships are represented by dark red, while the absence of correlations is shown in white. Missing correlations are indicated by blank areas, reflecting data sparsity in the real dataset. Notably, high correlations are observed among certain variables, such as WO and DY (r = 0.82) and DP and AX (r = 0.82), suggesting strong co-occurrence patterns between these symptom pairs. The uniformity of relationships across several pairs, particularly among DY, DP, AX, IB, and NFal, indicates a consistent and possibly systematic association within the real dataset.

In stark contrast, the synthetic dataset (Figure 2) exhibited predominantly weak correlations, mostly between -0.10 and +0.10, with few reaching statistical significance (e.g., DY and DP, r = 0.106, p = 0.018).

**Figure 2.**
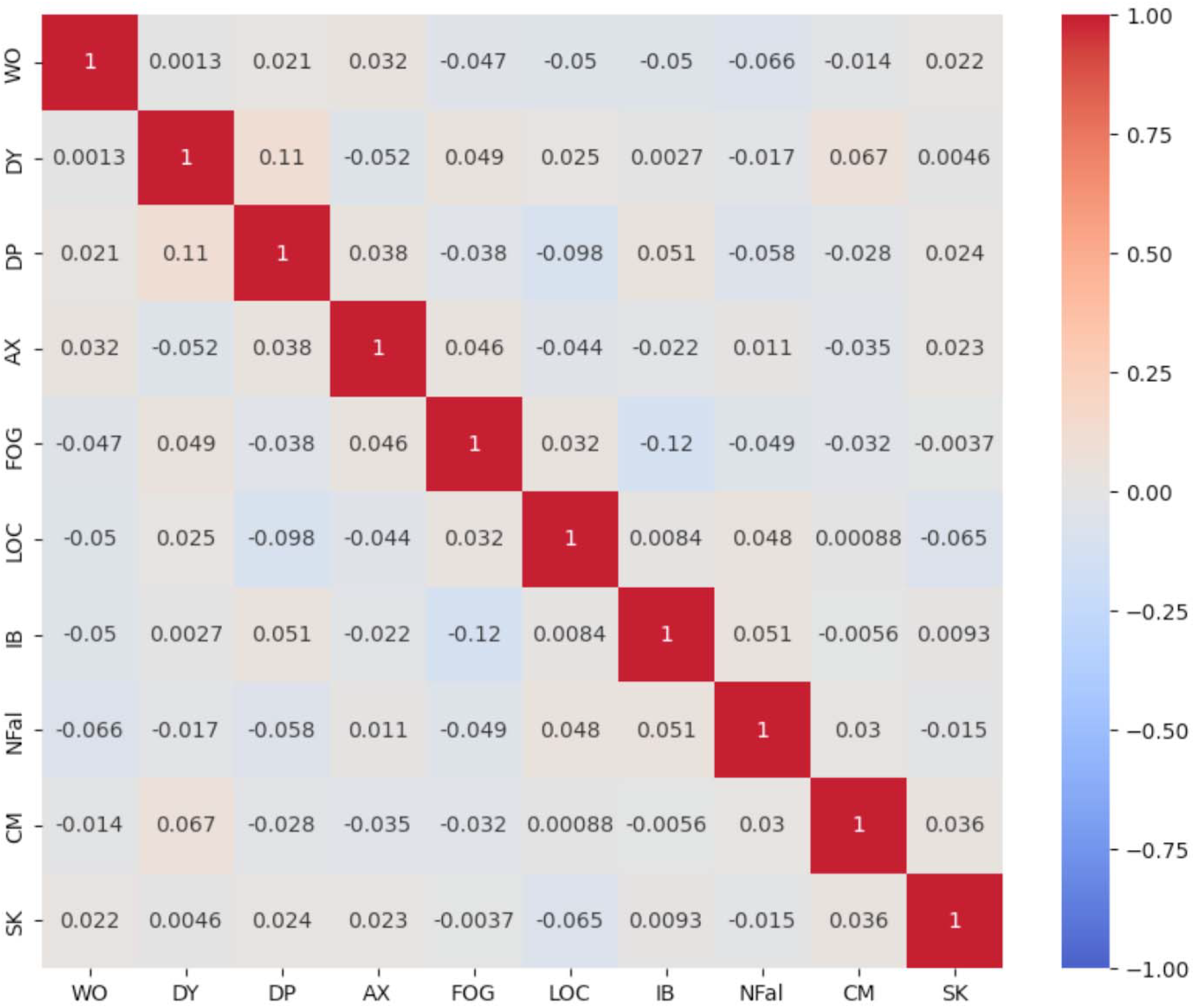
Spearman correlation matrix for symptom frequency variables in the synthetic dataset. This heatmap illustrates pairwise Spearman correlation coefficients (r) among ten symptom variables (WO, DY, DP, AX, FOG, LOC, IB, NFal, CM, SK), where coefficients range from -1 to 1. Positive correlations are represented in shades of red, indicating a direct relationship between variables, while negative correlations are shown in shades of blue, reflecting an inverse relationship. Correlations close to zero, shown in white, suggest little to no monotonic association. Diagonal values (r = 1) reflect perfect correlations of each variable with itself. The matrix highlights weak overall relationships between most variables, with coefficients predominantly clustered around zero. For instance, the correlation between DY and DP (r = 0.11) represents a weak positive relationship, whereas FOG and IB exhibit a small negative association (r = -0.12). This visualization provides an overview of the limited interdependence between symptoms within the synthetic dataset, which may indicate low co-occurrence patterns or reflect the independent nature of these variables in the simulated data.

Direct comparison of the flattened correlation vectors revealed substantial divergence: the Mean Absolute Difference (MAD) was 0.87, and the Root Mean Squared Difference (RMSD) was 0.88. The correlation of correlations was slightly negative (r = -0.044). This significant divergence indicates that GPT-4o, using only internal knowledge, failed to replicate the magnitude and pattern of inter-variable associations (symptom co-occurrence) observed in the real patient data.

### Thematic Analysis

Thematic analysis revealed distinct narrative structures and content. Synthetic data clustered around themes of **Emotional Coping** (e.g., “Felt anxious about falling, so stayed close to home”) and **Physical Adaptation** (e.g., “Walked around the park, but had to sit down often”). These coalesced into overarching themes of **Coping and Resilience** and **Adapting to Physical Limitations**, characterized by generalized emotional tones and pragmatic, yet impersonal, descriptions.

Real data clustering yielded themes of **Quantified Monitoring** (e.g., “Test 1 bad / Test 2 ok / Test 3 ok”, “1hr walk …no music”) and **Daily Reflections** (e.g., “Short 25 min walk; bad knee prevented more”). These formed overarching themes of **Structured Health Monitoring** and **Everyday Experiences**, emphasizing factual, systematic recording and specific situational details over emotional elaboration. A visual representation of these thematic structures is shown in Figure 3.

**Figure 3.**
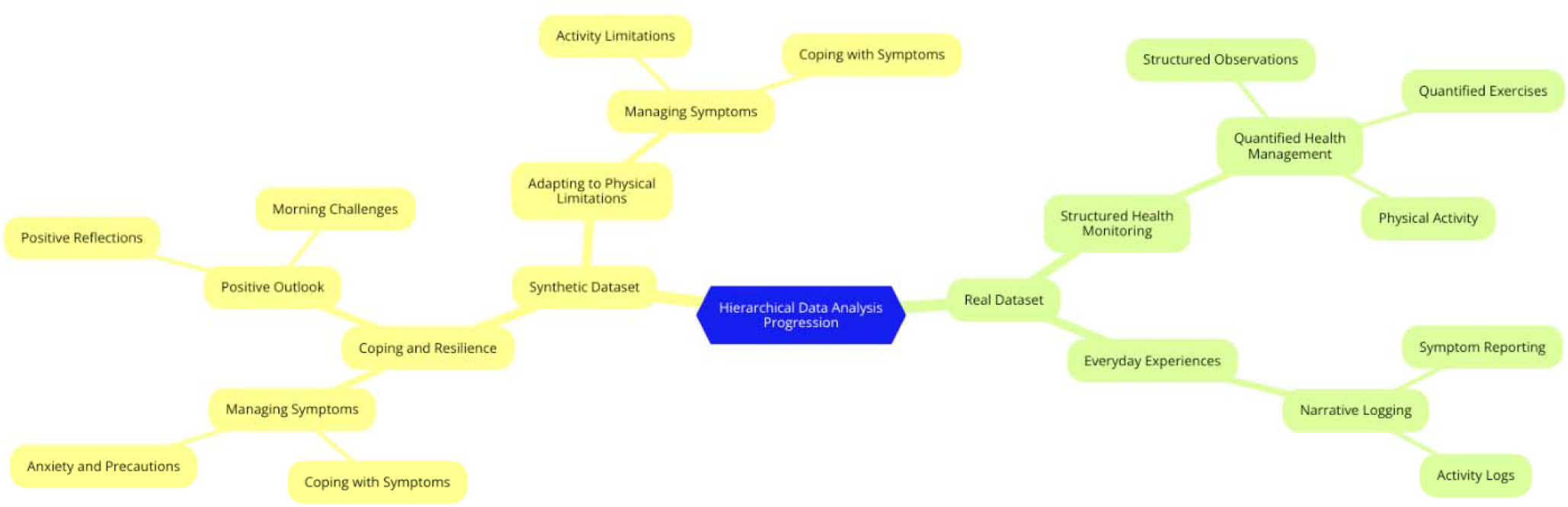
Hierarchical progression of selective themes, axial categories, and clusters for synthetic and real datasets. The diagram illustrates the relationships between high-level selective themes and their corresponding axial categories, which further branch into specific clusters. For the synthetic dataset, the selective themes are “Coping and Resilience” and “Adapting to Physical Limitations,” while for the real dataset, they are “Structured Health Monitoring” and “Everyday Experiences.” Each axial category connects to its respective clusters, highlighting the thematic structure and underlying data narratives.

### Narrative Analysis

Narrative analysis highlighted differences in storytelling. Synthetic entries were concise and general (e.g., “A little shaky today, but managed to get through”), lacking specific temporal or contextual details, aiming for universality but sacrificing richness. Real entries demonstrated greater linguistic complexity and specificity (e.g., “Excellent off walk 0615 to 0715 I was free from all toe cur and foot stiffness…”), often including causal links, reflections on stressors (e.g., “did not have my dog with me which caused increase stress”), and learning moments (e.g., “keep stress down, and ambulosono works for off walks”). Real entries also frequently featured structured observations (e.g., “Deep Water Workout -1 hour,” “Yoga - 1 hour”).

### Quantitative Content Analysis

Quantitative linguistic analysis validated qualitative observations. Word diversity was significantly lower in synthetic data (0.071) compared to real data (2.51), indicating more repetitive phrasing synthetically versus richer language in real entries. Average sentence length was slightly higher in synthetic data (9.52 words) than real data (7.94 words). Emotional terms (“anxious,” “trouble,” “hopeful”) appeared in 4.66% of synthetic entries but were absent in the analyzed real entries, aligning with the empathetic but potentially formulaic tone of synthetic data versus the factual style of real data. Passive voice constructions were more prevalent in synthetic data (1.95%) compared to real data (0.94%), suggesting a less agentic narrative style in synthetic entries compared to the active voice emphasizing patient agency in real diaries.

## Discussion

This study investigated GPT-4o’s capacity to generate synthetic Parkinson’s disease diary entries using only its internal knowledge, comparing the output to real patient data across thematic, narrative, and statistical dimensions. While GPT-4o replicated overall symptom prevalence distributions and some high-level themes, significant limitations emerged in capturing narrative richness, semantic depth, specific contextual details, and crucial inter-variable relationships found in real-world clinical data.

## Interpretation of Results

### Thematic Fidelity and Generalizability

Thematic analysis underscored a divergence in focus: synthetic entries emphasized generalized **Coping and Resilience**and **Adapting to Physical Limitations**, while real entries prioritized **Structured Health Monitoring** and specific **Everyday Experiences**. This suggests GPT-4o can synthesize broadly relevant, impersonal narratives reflecting common PD challenges but lacks the granularity of real patient experiences, which often include actionable insights tied to specific times, patterns, and causal links. The generalized emotional tone in synthetic data might stem from GPT-4o’s inherent design for broad applicability, potentially limiting its ability to mimic nuanced individual perspectives authentically.

### Narrative Structure: Strengths and Limitations

Narrative analysis confirmed the simplicity and universality of synthetic entries (e.g., “A little shaky today, but managed to get through”). While potentially useful for creating standardized datasets, this approach omits the complex interplay between symptoms, context, and behavior evident in real narratives that included detailed temporal markers, reflections, and learning moments (e.g., “Keep stress down, and ambulosono works for off walks”). This limitation suggests that while synthetic data offers coherence, it underrepresents the variability and richness vital for exploratory qualitative research, potentially oversimplifying patient behaviors and coping strategies.

### Linguistic Complexity and Implications for Usability

The lower word diversity (0.071 vs. 2.51) and higher passive voice usage (1.95% vs. 0.94%) in synthetic data suggest a trade-off between standardization and richness. The presence of emotional terms in synthetic, but not real, data highlights an empathetic yet potentially formulaic tone. This emotional accessibility might suit patient-facing applications but could reduce credibility in clinical research contexts demanding objective, precise data. The simpler, more passive structure contrasts with the active patient voice often seen in real diaries.

### Statistical Fidelity: Symptom Prevalence vs. Inter-Variable Relationships

A key finding is the dichotomy in statistical replication. GPT-4o accurately reproduced the *prevalence* of individual symptoms (Table 3). However, it failed dramatically to replicate the *relationships* between symptoms (Figures 1 & 2, MAD=0.87, RMSD=0.88). Real data showed strong correlations between certain symptoms, reflecting clinical co-occurrence patterns.

Synthetic data showed almost no correlation structure. This indicates that while GPT-4o’s internal knowledge contains information about *individual* symptom frequencies in PD, it lacks or cannot effectively utilize information about how these symptoms typically *interact* or co-occur in real patients. This is a critical limitation for research relying on understanding symptom clusters or complex interactions.

### Broader Implications and Critical Evaluation

These findings highlight the dual-edged nature of synthetic data generated solely from an LLM’s internal knowledge. Consistency, scalability, and accurate symptom prevalence distributions offer advantages for hypothesis generation, preliminary analyses, or addressing data scarcity.

However, reduced granularity, narrative simplicity, lack of correlation structure, and potential overgeneralization limit applicability for nuanced, patient-specific research or studies requiring high fidelity to real-world clinical patterns. The reliance on a single example entry and demographics also risks embedding input data limitations or biases into the output.

### Strengths and Limitations of Our Study

This study provides valuable initial insights into GPT-4o’s capabilities and limitations using internal knowledge for PD diary generation. A key strength is the multi-faceted evaluation framework combining qualitative and quantitative methods. However, limitations include the small sample size (n=57) of the real dataset, which may not fully represent broader PD population variability. Furthermore, this is a case study using a single synthetic dataset generated from one prompt structure; it doesn’t assess the consistency or variability of GPT-4o’s output across different prompts or generation runs. The findings are specific to using GPT-4o *without* fine-tuning on domain-specific diary data; performance might differ significantly with such training.

### Comparisons to Previous Research

Our findings align with research showing LLMs can generate statistically plausible synthetic healthcare data for some parameters [26] but struggle with capturing nuanced context and narrative richness compared to human-authored text [4, 13]. The successful replication of symptom prevalence echoes studies using GPT for generating standardized datasets [26], while the failure to capture correlations and narrative depth aligns with observations of AI generating context-poor or overly general content when not adequately trained or prompted [11, 23]. The linguistic simplicity observed contrasts with the recognized importance of narrative richness in patient diaries [4]. The struggle to replicate detailed narratives when not trained on specific diary data supports calls for context-rich training datasets [23].

### Generative Capabilities of GPT-4o: Strengths and Constraints

GPT-4o’s transformer architecture enables pattern recognition and probabilistic text generation, reflected in the structural and thematic consistency of the synthetic data and accurate symptom prevalence replication [2]. Its scalability addresses sample size issues [1]. However, its probabilistic nature favors generalized patterns over specific, context-dependent nuances, likely explaining the weak correlation structure and less granular symptom combinations (e.g., including BED/Fal predictions despite absence in the real sample subset, possibly reflecting broader internal knowledge not constrained by the specific input sample). This tendency towards plausible generalization over strict adherence to empirical patterns has been noted elsewhere [25, 29].

### Narrative Simplification and Emotional Generalization

The model’s optimization for coherence and fluency likely drives the emotionally generalized, simplified narratives [29]. Reliance on probabilistic word prediction based on vast but potentially non-specific training data can lead to omitting idiosyncratic details crucial to authentic patient narratives. Unlike real diaries integrating temporality and causality, GPT-4o defaulted to simpler structures, prioritizing general applicability over deep narrative coherence tailored to individual contexts [29].

### Statistical Variability and Overgeneralization

Differences in descriptive statistics (higher means, uniform IQRs, broader ranges in synthetic data) may also stem from GPT-4o balancing prompt specifications with its internal knowledge to ensure plausible variability [29]. This risks overgeneralization, where synthetic data might overrepresent certain combinations or exceed real-world variability, a tendency observed in other GPT applications [29].

### Potential Explanations for the Results: Training Data and Prompt Design

GPT-4o’s general training corpus likely lacks the specific distributional nuances and narrative styles of PD diaries, hindering specificity [11]. The model may default to common patterns in its training, reducing fidelity to rare phenomena. The prompt, using only a single example and demographic distribution, likely constrained the model’s ability to replicate the full variability and complexity of the real dataset [14]. While guiding structural replication, it may not have conveyed the necessary depth of context.

### Generalization Bias

As a model optimized for generalization, GPT-4o prioritizes broad applicability, which, while useful for scalability, can compromise reproduction of nuanced patterns intrinsic to specific real-world data like patient diaries [17].

### Implications and Future Directions of Generative AI in Clinical Research

This study highlights both the transformative potential and the inherent limitations of generative AI like GPT-4o in clinical research. Scalability and thematic consistency offer tools for addressing small sample sizes or enabling exploratory analyses. However, reliance on probabilistic synthesis without sufficient domain grounding risks overgeneralization and narrative simplification, limiting use where high fidelity to individual-level data is crucial.

To maximize the clinical relevance and applicability of synthetic data generated by AI, several strategic directions warrant consideration. Future research should focus on **iterative prompt refinement**, incorporating multiple, diverse sample entries and specifying greater contextual detail to guide models toward richer, more personalized outputs. Investigating **hybrid approaches** that strategically combine synthetic data with real-world datasets could offer a way to balance scalability with authenticity, ensuring more robust model training and validation.

**Domain-specific fine-tuning** presents another promising avenue, tailoring generative AI models to specific clinical domains by training them on high-quality, representative annotated datasets (such as actual PD diaries, where ethically feasible and available). Furthermore, methodological rigor requires careful **consistency evaluation**, assessing the variability of outputs across different generation runs and prompt designs. Extending research beyond single case studies to include **cross-context validation**by evaluating applicability in other chronic conditions, like Alzheimer’s disease or rheumatoid arthritis, will be essential to understand generalizability.

Crucially, **longitudinal validation** studies are needed to determine whether AI-generated datasets can accurately model temporal dynamics such as symptom progression and treatment responses over time. Finally, exploring methods for the explicit **integration of domain knowledge**, such as injecting known clinical symptom correlations or pathways into the generation process, could help bridge the gap between generalized outputs and clinically nuanced realities. By pursuing these directions, generative AI models can be refined into more reliable and ethical tools for advancing clinical research, improving patient outcomes, and complementing traditional data collection methods.

## Conclusion

This case study demonstrates GPT-4o’s potential for generating statistically plausible synthetic Parkinson’s diary entries regarding overall symptom frequencies when relying solely on its internal knowledge, offering a scalable, privacy-preserving approach. However, this method significantly limits its ability to capture the narrative richness, contextual depth, and crucial inter-variable correlation structures present in real patient experiences. While potentially valuable for preliminary exploration or addressing data scarcity in low-fidelity scenarios, rigorous validation and refinement, likely involving domain-specific data or knowledge integration, are necessary before such AI-generated data can be reliably used for research requiring high narrative fidelity or accurate representation of complex clinical patterns in Parkinson’s disease.

## Data Availability

All data produced in the present study are available upon reasonable request to the authors

## Acknowledgements

This research was supported by CIHR, Alberta Innovation and Ministry of Mental Health and University of Calgary Endowment Fund

